# Effectiveness of tele-exercise on muscle function and physical performance in older adults for preventing sarcopenia: A protocol for systematic review

**DOI:** 10.1101/2024.03.06.24303856

**Authors:** Ya Shi, Emma Stanmore, Lisa McGarrigle, Chris Todd

## Abstract

**Introduction:** Sarcopenia is characterized by the progressive weakening of muscle function that occurs with age. This condition frequently leads to frailty, disability, and even death. Research on sarcopenia prevention is growing. Tele-exercise intervention is increasingly gaining attention in this field, with the rapid advancement of the Internet and the influence of the COVID-19. However, there is a lack of empirical support for its effectiveness. Our study aims to assess the effect of tele-exercise on sarcopenia in older persons, specifically focusing on its ability to improve muscle strength, muscle mass and physical performance.

**Methods and analysis:** Searching will be performed in the following eleven databases (Medline, Embase, Cochrane Central Register of Controlled Trials, CINAHL, PsycINFO, WOS, Scopus, CBM, CNKI, WANFANG, VIP) for published trials and two trial registries (Clinicaltrials.gov and the WHO International Clinical Trials Registry Platform) for unpublished trials. Google Scholar will be utilized to find grey literatures. The criterion of inclusion will be clinical trials involving tele-exercise interventions in older adults (≥ 60y) diagnosed with sarcopenia (possible, confirmed, or severe sarcopenia). For data synthesis, we will utilize a summary table to show the major characteristics of selected trials and a summary graph to demonstrate the risk of bias using RoB 2 in each trial, which will be further discussed in a narrative synthesis. The possibility of meta-analysis for quantitative data will be assessed according to the homogeneity analysis of the trials, using the methods of fixed or random effects model. If meta-analysis is possible, subgroup analysis and sensitivity analysis will be performed as well. Publication bias will be assessed through the use of the funnel plot and Egger’s linear regression test when an adequate number of trials are available. Finally, the GRADE approach will be used to classify the certainty of evidence body into four categories (high, moderate, low, and very low).

**Ethics and dissemination:** The findings of the systematic review will be shared through publishing in a peer-reviewed journal and presentation at appropriate conferences. Since we will not be utilizing specific patient data, ethical approval is unnecessary.

**PROSPERO registration number:** CRD42024516930

**Strengths and limitations of this study:**

- This will be the first systematic review on tele-exercise for sarcopenia prevention in older adults.
- This review will seek to determine the duration, frequency, intensity and type of tele-exercise that is most appropriate for preventing and treating sarcopenia.
- The results may fill the gap pertaining to accurate tele-exercise prescription in older adults with sarcopenia.
- This review will adhere to the PRISMA standards for conducting and reporting systematic reviews and meta-analyses in order to reduce bias.
- High heterogeneity may exist due to the different diagnostic criteria and thresholds for sarcopenia.

## BACKGROUND

Sarcopenia is a muscle weakness or muscle failure caused by adverse muscle changes that accumulate over a lifetime and has a very high prevalence among older individuals, according to the recent European consensus (the European Working Group on Sarcopenia in Older People 2, EWGSOP 2) [1]. A systematic review examined nine studies and revealed that the sarcopenia prevalence in older adults was 17.7% by referring EWGSOP1 and 11% by referring EWGSOP2 [2]. The prevalence of sarcopenia is also considerably elevated among the older population in Asian. A meta-analysis indicated that the overall prevalence of sarcopenia in older Chinese men and women was 18.0% and 16.4%, respectively [3]. A 5.8 year prospective study of 1851 Japanese older adults revealed that the sarcopenia prevalence was 11.5% in men and 16.7% in women [4]. Another meta-analysis analysed three studies and found that the total prevalence of sarcopenia in older Korea males was 14.9%, while it was 11.4% in older Korean females [5]. In general, the global prevalence of sarcopenia in older people aged 60 years and older ranges from 10% to 27% based on different classifications and cut-off points for sarcopenia diagnoses [6].

Providing optimal care for older individuals with sarcopenia is crucial due to the significant personal, societal, and economic difficulties associated with the untreated and deteriorated condition [7]. First, sarcopenia may affect the health of older population. Sarcopenia is associated with different acute and chronic diseases in older patients, such as hypertension [8], diabetes mellitus [9], coronary artery disease [10], heart failure [11], asthma [12], chronic kidney disease [13], etc. In addition, sarcopenia is also linked to an increased vulnerability to falls and fractures [14], diminished capacity to perform daily activities [15,16], heightened risk of mobility impairments [17], greater requirement for long-term care placement [18], and ultimately contributes to a deterioration in quality of life [19], and even higher mortality rates [20]. Second, sarcopenia imposes a serious economic burden on society and families. Evidence from a study involving 1358 community-dwelling older adults revealed that sarcopenia was linked to the likelihood of hospital admission [21]. A prospective cohort study found that sarcopenia resulted in higher hospitalization costs in convalescent rehabilitation units among older patients with sarcopenia than those without [22]. Hence, it is imperative to prioritize early detection, prevention, and treatment of sarcopenia.

Exercise is the most studied and critical method at present among different types of non-pharmacological intervention for preventing sarcopenia. Our previous scoping review summarised 59 studies that focused on non-pharmacological interventions for sarcopenia prevention in community-dwelling older adults [23]. The review revealed that the interventions that included exercise component accounted for a significantly higher percentage (52.8%) compared to those that included nutrition (34.5%), health education (15.5%), and traditional Chinese medicine (2.1%). A systematic review and meta-analysis evaluated 22 studies and found that exercise treatment had overall significant positive effects on muscle strength and physical performance but not on muscle mass in older adults with sarcopenia [24]. Another meta-analysis included seven studies based on the EWGSOP criteria but substantiated comparable findings [25]. An additional network meta-analysis further revealed that both exercise in isolation and the combination of exercise and nutrition yielded favourable outcomes in terms of muscle strength and physical performance among sarcopenic older adults [26]. Moreover, a systematic review of systematic reviews examined the effects of different modalities of exercise intervention on older individuals with sarcopenia. The findings indicated that resistance training was better for both muscle strength and skeletal muscle mass, while mixed modalities (resistance training and non-resistance training) was better for physical performance [27]. Furthermore, Hurst et al. [28] highly recommended resistance exercise as the first-line treatment for counteracting the deleterious consequences of sarcopenia in older individuals.

Due to the rapid advancement of the Internet and communication technologies, as well as the influence of the COVID-19 pandemic, tele-exercise programs are gaining increasing interest in the field of sarcopenia prevention. Tele-exercise is a component of telehealth, which is generally conducted online through the use of internet-connected devices such as computers, tablets, and smartphones. For example, Chan et al.[29] designed an online exercise programme via Zoom for older people with possible sarcopenia or at risk of fall. Tuan et al. [30] created an intervention using Nintendo Switch RingFit Adventure to explore the clinical effectiveness of exergame-based exercise on muscle function and physical performance among older people. Indeed, certain studies have successfully obtained some results in this research area. For instance, Wang et al. [31] explored the effectiveness of an app on sarcopenia prevention in older adults and found that skeletal muscle mass after the intervention was higher in the comprehensive (nutrition plus exercise) and nutrition groups than in the control and exercise groups. Hong et al. [32] developed a real-time tele-exercise intervention through Skype ™ for community-dwelling older adults with sarcopenia and indicated that this form had beneficial effects on factors related to sarcopenia such as total-body skeletal muscle mass, lower limb muscle mass, and the chair sit-and-reach scores. Besides, Yamada et al. [33] found a six-month mail-based intervention (exercise alone or plus nutrition) for sarcopenia prevention significantly improved anabolic hormone levels and skeletal muscle mass index in community-dwelling older adults.

However, regarding tele-exercise in sarcopenia prevention field, we were unable to identify any literature review specifically addressing its real effects on muscle function and physical performance of older individuals. There remain unresolved questions and paradoxes that require resolution. For example, the meta-analysis has already established that the conventional form of exercise did not have any beneficial impact on muscle mass [24, 25], but the subsequent studies have indicated that tele-exercise did have a good effect on this index [32, 33]. Besides, as we mentioned above, there exist different types of remote devices for older people to prevent sarcopenia, such as Zoom, Skype™, email, and even self-developed app [29-33]. Apart from tele-exercise, the intervention for preventing sarcopenia in older population also encompasses tele-nutrition, as well as the combination of tele-exercise with tele-nutrition and other approaches [31, 33].

Therefore, our primary aim is to conduct a systematic review to address the following research questions: 1) What are the influences of tele-exercise on sarcopenic indices (muscle mass, muscle strength, and physical performance) in older adults before and after intervention? 2) What is the comparative efficacy of tele-exercise in preventing sarcopenia, as opposed to tele-nutrition, tele-exercise and tele-nutrition combined, or conventional intervention? 3) Which type of remote devices offers optimal benefits in older adults for sarcopenia prevention and treatment? The study findings will consolidate the evidence to address these inquiries, potentially facilitating the utilization of tele-exercise as a non-pharmacological or supplementary intervention for preventing sarcopenia among older people, establishing a basis for future research, and providing significant insights for researchers in the corresponding discipline.

## METHODS AND ANALYSIS

### Reporting

The protocol follows the guidelines of Preferred Reporting Items for Systematic Review and Meta-Analyses Protocols (PRISMA-P, as shown in online supplemental material S1) [34, 35] and the recommendations for systematic reviews involving older adults by Shenkin et al. [36], to guarantee comprehensive reporting and execution. The review methodology was already preregistered on the International Prospective Register of Systematic Reviews (PROSPERO) with registration number CRD420245 16930 [37].

### Eligibility criteria

We utilized the “PICO” principle [38] to establish the eligibility criteria for this study and will choose primary studies based on the criteria below.

### Inclusion criteria

#### 1. P-Population

Older adults with sarcopenia will be considered. In terms of age definition of an older person, although it is not uniform to some extent around the world according to different conditions by different countries (e.g.≥50 in Africa, ≥60 in United Nations and China, ≥65 in western countries, ≥75 in Japan), the ages of 60 and 65 years are often used [39-41]. We will include papers with a study population ≥ 60 years old or an average age ≥ 60 years old, so as to incorporate as many references as possible. Various definitions and diagnostic criteria for sarcopenia exist [1, 42-44], hence this study will utilize criteria and cut-off points established in previous research that conducted musculoskeletal measurements and select studies that recruited older adults with reduced muscle strength and mass. Meanwhile, according to the latest international classification standard for sarcopenia [1, 44], studies on the three categories including possible, confirmed and severe sarcopenia will all be included.

#### 2. I-Intervention

We will include studies with any form of tele-exercise lasting at least 4 weeks [45]. First, the prefix “tele” indicates that the exercise is conducted online with the assistance of internet-enabled devices like computers, tablets, and smartphones. Second, “exercise” encompasses a range of training modalities like resistance training, aerobic training, balance training, and more.

#### 3. C-Comparators

The groups performing no tele-exercise (i.e. traditional nutrition/health education/ usual care without Internet devices, tele-nutrition/health education/usual care) or a sham tele-exercise intervention will be considered as comparators.

#### 4. O-Outcomes

Common measurements in sarcopenia research will be taken into account and must be measured both before and after the intervention. We utilized the classification method established in the earlier scoping review [23] to categorize outcome measures.

##### Main outcomes: 1)

For muscle strength assessment, four types is grouped based on parts of the body being measured, including hand grip strength, back strength, upper limb extension strength and lower limb flexion strength. 2) For muscle mass, three types is grouped also based on parts of the body being measured, containing appendicular, trunk and whole-body muscle mass. 3) For physical performance, four types is grouped based on the assessed contents, including gait speed (e.g.6 minutes-walk test, 10-m walk test and 4-m walk test), functional ability (e.g. 4-step stair climb performance, timed-up-and-go test, 8-foot up and go test and 5 times sit-to-stand test), balance ability (e.g. standing on one foot, Berg balance scale and sensory motor control), comprehensive physical performance (e.g. Short Physical Performance Battery, Barthel index, Activities of Daily Living and Physical Activity Scale for the Elderly).

##### Other outcomes: 1)

For other body composition, three types is grouped, including obesity measurements (e.g. body mass index and body fat mass), bone mass/mineral (e.g. bone mass and bone mineral content) and body circumference (calf circumference and waist circumference). 2) For general health status, this type contains the outcomes assessed by comprehensive scales reflecting quality of life, fall risk, sleep quality and so on (e.g. Sarcopenia Quality of Life Scale, EQ-5D-5L Quality of Life Questionnaire, 25-question Geriatric Locomotive Functional Scale and Modified Falls Efficacy Scale). 3) For nutrition state, this type includes single or multicomponent nutrition assessment, like Mini-Nutritional Assessment Questionnaire (MNA), Energy Intake Assessment, Protein Intake Assessment and so on.

#### 5. Study design

Randomised controlled trials and blind or open clinical trials [(quasi-) RCTs (parallel and crossover)] will be selected.

### Exclusion criteria

1. Unfinished or ongoing studies or study protocols will be excluded.
2. Researches on sarcopenia concomitant with another disease (i.e. cancer, cachexia, obesity, hemodialysis, neurologic disease) will be excluded.
3. Studies focusing on animals, genetics or biochemistry will not be considered.
4. Qualitative researches, observational studies (i.e. cohort, cross-sectional or case-control study), reviews (i.e. systematic review, meta-analysis, scoping review, narrative review), opinion/perspective articles, conference abstracts, editorials, case reports and comments will be excluded.
5. Publications meeting the inclusion criteria but with unavailable results even after consulting the authors will be excluded.

### Database search

Our search will be conducted in eleven databases [Medline, Embase, Cochrane Central Register of Controlled Trials, Cumulated Index to Nursing and Allied Health Literature (CINAHL), Psychological Information (PsycINFO), Web of Science (WOS), Scopus, Chinese Biomedical Literature Database (CBM), Chinese National Knowledge Infrastructure (CNKI), Wan Fang Database (WANFANG), Chinese Science and Technology Periodical Database (VIP)] for published trials and two trial registries (Clinicaltrials.gov and the WHO International Clinical Trials Registry Platform) for unpublished trials. Besides, Google Scholar will be utilized to find grey literatures. Furthermore, the references of the chosen articles will also be checked to complement the search and ensure thorough coverage of the literature.

Search strategy after consultation with a professional librarian focuses on population, intervention and study design, without limitations on language and publication period. Searching example for WOS is shown in Table 1. For other databases, the search strategy will be adjusted based on the specific requirements of each database. Searches in all selected databases will be formally carried out in April 2024. A copy of the search strategies for these databases and preliminary search results will also be saved.

**Table 1.** WEB OF SCIENCE search strategy

| NUM | SEARCH STRATEGY | RESULT |
| --- | --- | --- |
| 1 | TS=(older adult* or older person or elder* or geriatric* or senior* or veteran* or aged or aged, 80 over or oldest old or age, eld* or nonagenarian* or octogenarian* or centenarian*) |  |
| 2 | TS=(sarcopeni* or muscular atrophy or muscle weakness or muscle loss or muscle depletion or muscle reduction or muscle wasting or loss of muscle or low muscle mass or low muscle strength) |  |
| 3 | TS=(tele* or remote or distance or ehealth or e-health or digital health or mhealth or m-health or mobile health or web* or Internet or online or computer* or tablet* or smartphone* or app* or mail* or video* or electronic or social media or Twitter or Facebook or Instagram or YouTube or WhatsApp or Microsoft Teams or Zoom or TikTok or WeChat or Weibo or QQ) |  |
| 4 | TS=(physical activit* or exercise* or sport* or training or coaching) |  |
| 5 | TS=(randomized controlled trial* or RCT* or controlled trial* or clinical trial*) |  |
| 6 | #1 AND #2 AND #3 AND #4 AND #5 |  |
| 7 | TI=(qualitative stud* or cross-sectional stud* or cohort stud* or case-control stud* or review* or meta-analysis or protocol* or conference* or case report* or comment*) |  |
| 8 | (#6) NOT #7 |  |

### Review process

The entire review process consists of three distinct parts: searching, integrating, and selecting stages, which will be finalised by the end of June 2024. During searching stage, two researchers (YS and JYD) will independently search each database simultaneously according to the corresponding searching strategy, then comparing the amount of references in each database. If there is any disagreement, a third member of the research team (CT, LMcG or ES) will check again. During integrating stage, all references will be integrated into Endnote software to remove duplicates and then will be transferred to Rayyan software for screening [46]. During selecting stage, two independent researchers (YS and JYD) will first screen the literatures using the title/abstract/keywords and then using full-text to determine the suitability of each article, based on the inclusion and exclusion criteria mentioned above. Disagreements over selection will be addressed and settled through consensus with a third member of the research team (CT, LMcG, or ES). Finally, eligible articles will be included in the systematic review. According to PRISMA-P recommendations, we will prepare a flowchart (as shown in online supplemental material S2) with necessary information about selection process, including the total number of references in different stages, and specific reasons for inclusion and exclusion.

### Data extraction

Two researchers (YS and JYD) will independently conduct data extraction, with any differences being handled by a third researcher (CT, LMcG, or ES). Targeted data will be extracted from an article using a standardized form created by our research team. We will consider the following aspects: 1) research characteristics: author, publication year, study setting, study design, sample size, diagnostic criteria for sarcopenia; 2) population characteristics: age range (average age), gender/sex, co-morbidity, cognition, cultural background; 3) intervention characteristics: intervention type (i.e. exercise, nutrition, health education), delivery tool (i.e. web, app, mail, social media), intervention dose (i.e. duration, frequency, intensity), follow-up period, compliance, drop-out, adverse events related to intervention; 4) outcome characteristics: baseline and follow-up values of six categories as mentioned above, including muscle strength, muscle mass, physical performance, other body composition, general health status, and nutrition state. Two researchers (YS and JYD) will test the form on three articles before its official implementation. If pertinent information is absent, we will contact the corresponding author/s twice at weekly intervals. We intend to finalise the data extraction process at the end of July 2024.

### Risk of bias in individual trials

Two researchers (YS and JYD) will evaluate the risk of bias separately using the revised Cochrane risk-of-bias tool for randomized trials (RoB 2) [47] after assessing three pilot trials, without being blinded to the authors and journal of the primary studies. The researchers will select the corresponding version of the RoB 2 for each trial according to different study designs (individually-randomized parallel-group trials, cluster-randomized trials, and crossover trials). The tool is structured into five domains: 1) bias arising from the randomization process; 2) bias due to deviations from intended interventions; 3) bias due to missing outcome data; 4) bias in measurement of the outcome; 5) bias in selection of the reported result. Each domain will be assessed using specific algorithms based on responses to relevant signalling questions with five response options (yes, probably yes, probably no, no, no information). Domain-level risk of bias will then be classified as low risk, some concerns, or high risk. The overall risk of bias on study level will also be rated as low risk (all domains in low risk of bias), some concerns (at least one domain in some concerns but no domain in high risk of bias), or high risk (at least one domain in high risk of bias or multiple domains in some concerns). Discrepancies will be resolved through discussion between the two researchers (YS and JYD) and a third researcher if required (CT, LMcG, or ES). The final results will be displayed in a risk-of-bias graph, which will be completed by the end of September 2024.

### Data synthesis

Data synthesis mainly includes the integration of qualitative data and quantitative data. For qualitative analysis, we will utilize a summary table to display the main characteristics of each trial and a summary graph to illustrate the risk of bias in each trial, which will be then discussed in a narrative synthesis. For quantitative data outcomes, meta-analysis will be conducted using Review Manager (RevMan, version 5.4) software. Typically, at least two studies are necessary to conduct a meta-analysis [48].

### ·Effect sizes

Effect sizes will be used to determine the effect of two interventions on different variables. Effect sizes for dichotomous variables, such as negative health outcomes like mortality, will be presented as risk ratios (RR) or odds ratio (OR) with a 95% CI. Effect sizes for continuous variables, such as muscle strength, muscle mass, and gait speed, will be presented as weighted mean difference (WMD, if all trials utilize identical measurement tools and units) or standardized mean differences (SMD, if trials utilize diverse measurement instruments or distinct units) along with a 95% confidence interval (CI). If the median is displayed, the median and interquartile range (IQR) will be converted to mean and standard deviation (SD) using the statistical formula [49].

### Heterogeneity analysis

The analysis of heterogeneity can be divided into three main categories: statistical, clinical, and methodological heterogeneity. Statistical heterogeneity will be visualised in forest plots (with 95% CIs of effect sizes) and be evaluated using a chi-squared test (with *p* value and *I*^*2*^ index) [50]. The *p* value can show the heterogeneity with or without statistical significance. In addition, the *I*^*2*^ index ranges from 0 to 100% and reflects the level of heterogeneity, with the higher *I*^*2*^ index indicating the greater heterogeneity. If the selected trials are shown as homogeneous (*p* ≥ 0.10) or low heterogeneity (*I*^*2*^ < 50%), a fixed-effects (FE) model will be applied to estimate the data. Conversely, if the chosen trials exhibit statistically significant heterogeneity (*p* < 0.10) or substantial heterogeneity (*I*^*2*^ ≥ 50%), a random-effects (RE) model will be employed to combine the data.

### ·Subgroup analysis

Subgroup analysis is mostly utilized in two scenarios: 1) If statistical heterogeneity is significant, potential sources of clinical and methodological heterogeneity will be identified and analysed using subgroup analysis based on clinical and scientific experiences. 2) If statistical heterogeneity is not significant, subgroup analysis is primarily used to examine the correlation between subgroup factors and outcomes. Depending on the situation, subgroups may be divided by age (i.e. 60-69y, 70-79y, and ≥ 80y), gender/sex, three categories of sarcopenia (possible, confirmed, or severe sarcopenia), different diagnostic criteria for sarcopenia (i.e. AWGS 2014/2019, EWGSOP 2010/2019), different type of delivery tool (i.e. web, app, mail, social media), different modalities of tele-exercise (i.e. resistance training, aerobic training, balance training), intervention duration (i.e. ≤ 6 months, >6 months), comorbidities (i.e. with frailty, without frailty), and so on.

### Sensitivity analysis

Sensitivity analysis is applied to assess the robustness of the findings or conclusions derived from the primary meta-analysis of data in clinical trials [51]. We will conduct sensitivity analysis using the leave-one-out test in every primary meta-analysis of each outcome variable. The sensitivity analysis will cover all trials selected and use a one-by-one exclusion method before re-running the meta-analysis. If the point estimate of the combined effect size, after excluding a study, is beyond the 95% CI of the total combined effect size, it suggests that the study significantly influences the results; on the other hand, it suggests that the results are stable. The final results will be displayed in corresponding sensitive graphs.

### Publication bias

Publication bias will be evaluated by examining the symmetry of the funnel plot and conducting Egger’s linear regression test when at least 10 trials are included in meta-analysis [52]. Conclusions about publication bias may be uncertain due to the small number of studies for each outcome and the limited ability of these tests to detect publication bias. Additionally, Egger’s test may have some limitations when evaluating continuous outcomes [53]. We will use the Duvall and Tweedle trim-and-fill model to alter the effect estimates if there is an indication of publication bias [54]. Besides, reporting bias risk is minimized by requesting any pertinent results not expressly provided in research from the authors. If study authors do not respond, the review document will address the possibility of reporting bias. We will also analyze if the authors of the trials included have considered the effects of potential conflicts of interest and provided information on ethical approval [55].

### Certainty of the evidence

Grading of Recommendations, Assessment, Development and Evaluation (GRADE) approach will be used to assess the certainty of evidence [56, 57]. The GRADE categorizes the certainty for a body of evidence (rather than individual studies) into four categories (high, moderate, low, and very low) based on factors such as study design, the risk of bias, heterogeneity, indirectness, imprecision of study results, and publication bias. We intend to finalise the data synthesis process at the end of December 2024. The final review report will be generated according to the PRISMA standards after synthesizing and classifying the data as described.

## Data Availability

Data sharing not applicable as no data sets generated and/or analysed for this study.

## Patient and public involvement

No patients involved.

## Ethics and dissemination

Due to the nature of this study (systematic review), ethical considerations are not applicable, and ethical approval is unnecessary. All review findings will be shared extensively through peer-reviewed journals and conferences.

## Acknowledgements

The authors would like to thank: Ms. Claire Hodkinson, Medical Librarian at The University of Manchester Library, for her assistance with search strategy planning.

